# Embedding National Benchmarking Within Integrated Intermediate Care Pathways: A Quality Improvement Protocol Using BEPOP to Enhance Exercise Provision for Frail Older Adults in Cambridgeshire and Peterborough

**DOI:** 10.1101/2025.10.24.25338713

**Authors:** Ishmeet Singh, Barry Underwood, Alex Milne

## Abstract

**Background:** Frailty and sarcopenia are leading contributors to disability and dependence in older adults. Despite robust evidence for resistance-based exercise interventions, clinical practice remains inconsistent, with wide variation in assessment, prescription, and follow-up.

**Aim:** To embed the national BEPOP (Benchmarking Exercise Programmes for Older People) framework within CPFT’s Intermediate Care Service, integrating inpatient (Pathway 2) and community (Pathway 1) rehabilitation data to establish a baseline, implement targeted improvements, and evaluate outcomes post-intervention.

**Methods:** A pre–post quality improvement design will involve participation in BEPOP benchmarking audit across two different waves as a structured framework. Data for 20 older adults (≥65 years) with frailty or sarcopenia will be audited and shared with the BEPOP team. Data across six stages—baseline, assessment, delivery, review, post-intervention, and follow-up— will be collected via REDCap and benchmarked nationally by the BEPOP team. CPFT will evaluate local process fidelity, training impact, and documentation quality through a Plan– Do–Study–Act (PDSA) cycle spanning BEPOP Waves 3 and 4 (2025–2027).

**Measures:** Primary: proportion of patients receiving objective strength assessment and follow-up reassessment.

Secondary: exercise prescription fidelity, and clinician confidence.

**Anticipated Results:** Improved standardisation of assessment, enhanced cross-pathway collaboration, and strengthened integration of evidence-based exercise within routine frailty rehabilitation.

**Impact:** This initiative represents the first integration of BEPOP across both inpatient and community rehabilitation settings, offering a scalable model for embedding national benchmarking within local NHS services.

## Introduction

Frailty and sarcopenia are major geriatric syndromes linked with falls, hospitalisation, and loss of independence. Yet they remain under-recognised and sub optimally managed. Early detection and targeted resistance training can maintain mobility and independence, but implementation gaps persist. Less than half of older adults with sarcopenia and frailty receive structured resistance exercise, and objective strength assessments are inconsistently applied (1–3).

Within the National Health Service (NHS) of United Kingdom, rehabilitation practice varies widely across regions and care settings. This variation limits inter-service comparability, impedes evaluation, and constrains national learning.

The Benchmarking Exercise Programmes for Older People (BEPOP) project—coordinated by the National Institute for Health Research (NIHR) Newcastle Biomedical Research Centre, the British Geriatrics Society, and Allied Group for Interdisciplinary Leadership and Education (AGILE) which is Chartered Society of Physiotherapist’s Older People Special Interest Group—provides a standardised national audit to benchmark and enhance exercise prescription for older adults with frailty (4).

Aligning with NHS England’s Ageing Well and Intermediate Care frameworks, this project leverages BEPOP to embed a systematic benchmarking process across the integrated intermediate care pathways of Cambridgeshire and Peterborough NHS Foundation Trust (CPFT), enabling continuous improvement through iterative feedback cycles.

## Problem Statement

Exercise provision for frail older adults lacks consistent benchmarking across inpatient and community rehabilitation teams. Variation exists in:

- Use of standardised frailty and sarcopenia assessments
- Delivery and progression of resistance-training programmes
- Reassessment and documentation practices

This heterogeneity limits service evaluation and continuous improvement.

## Aim and Objectives

### Aim

To benchmark and improve exercise prescription for older adults with frailty or sarcopenia across CPFT’s Intermediate Care Pathways through integration of the BEPOP framework.

### Objectives

1. Establish a baseline of current assessment and exercise practices (Wave 3).
2. Identify variation and gaps through BEPOP national feedback.
3. Implement targeted improvement actions: staff training, standardised tools, and inter-pathway collaboration.
4. Re-evaluate practices post-intervention (Wave 4).
5. Embed BEPOP benchmarking into routine service review and quality improvement cycles.

## Rationale

Benchmarking quantifies variation and drives adoption of evidence-based improvement. Integrating BEPOP within CPFT’s Pathway 2 (inpatient units) and Pathway 1 (community teams) enables tracking of a single cohort across the six-week rehabilitation continuum— capturing the full trajectory from admission to reablement without altering length of stay or usual care.

This cross-pathway integration is novel nationally and represents a practical model for regional frailty systems.

## Methods

### Context

Intermediate care service of Cambridgeshire and Peterborough (Fig.1)

- **Intermediate Care Pathway 2 (Inpatient Rehabilitation):** four units across Cambridge, Peterborough, Ely and Wisbech.
- **Intermediate Care Pathway 1 (Community Rehabilitation):** teams covering Cambridge, Huntingdon, Peterborough and Fenland.

**Fig. 1.**
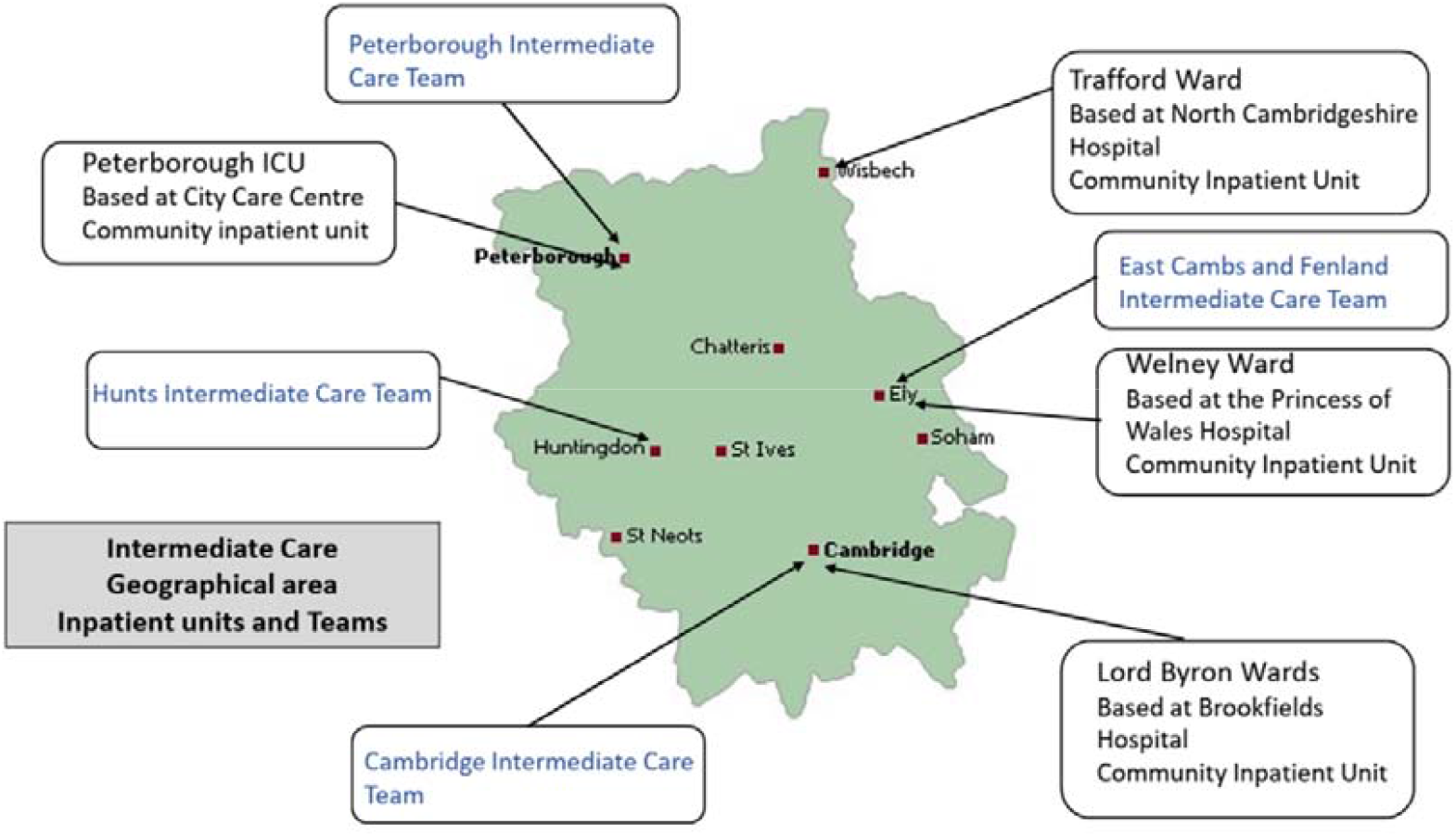
Intermediate Care Service map- Cambridgeshire and Peterborough

Together they serve urban and rural populations, ensuring inclusivity and representativeness.

### Intervention

Implementation of the BEPOP national audit framework within both pathways to establish baseline practice with BEPOP Wave 3 and guide targeted improvement actions leading to re-audit by participating in BEPOP Wave 4.

A Plan–Do–Study–Act (PDSA) cycle (5) will be implemented spanning BEPOP Wave 3 and Wave 4 (2025–2027).

#### Plan (Oct – Dec 2025)

- Collect baseline data via participation in Wave 3. De-identified data for 20 patients about baseline patient information, outcome measures used during initial assessments, frequency and intensity of intervention given, frequency and nature of review and progression and data from post intervention assessments and suggested follow up will be collected for 20 patients ≥65 years with frailty/sarcopenia.
- Submit anonymised data to BEPOP team for national benchmarking and receive feedback.

#### Do (Throughout 2026)

- Implement quality improvement actions informed by Wave 3 feedback:
  - Training on progressive resistance exercise.
  - Standardisation of frailty and strength assessments.
  - Enhanced documentation templates.
  - Inter-pathway feedback loops and AHP engagement.
  - Participate in BEPOP Wave 4 and contribute data.

#### Study (early 2027)

- Receive BEPOP Wave 4 feedback and analyse local CPFT process metrics:
  - Percentage of patients assessed and re-assessed
  - Programme progression rates
  - Staff confidence surveys
- Compare BEPOP Wave 3 and Wave 4 performance.

#### Act (ongoing post Wave 4-2027)

- Sustain improvements through local Standard operating procedure (SOP) revisions and continuous education.
- Disseminate learning internally and nationally via BEPOP and AGILE networks.

Each BEPOP wave constitutes an iterative feedback-driven learning cycle within this overarching PDSA framework.

### Measures

Primary process metrics:

- Proportion of patients receiving objective strength assessment at baseline.
- Proportion of patients receiving reassessment during the programme.

Secondary process metrics / service measures:

- Fidelity and consistency of exercise prescription (e.g., frequency, resistance components, progression).

Benchmarking:

- The BEPOP team will provide comparative benchmarking data nationally, which will inform Wave 4 improvement interventions.
- CPFT will use these reports to plan targeted quality improvement initiatives and re-audit post-intervention. Comparative analysis of Wave 3 and Wave 4 data will quantify changes in adherence to national standards.
- Qualitative data from staff will be thematically analysed.

## Data Analysis

- National benchmarking analyses undertaken by the BEPOP team.
- Local CPFT analysis will describe process metrics and qualitative themes from staff feedback using descriptive and thematic analysis.

### Governance and Ethics

Registered as CPFT Service Improvement Project. Caldicott Guardian approval obtained for data sharing.

As analysis involves anonymised routine care data, formal ethics review is not required under NHS HRA guidance.

### Patient and Public Involvement

Patients and the public were not directly involved in protocol design; findings will be disseminated through CPFT’s public engagement forums and annual Quality Report.

### Anticipated Outcomes and Impact

- Established CPFT baseline for exercise provision in frailty rehabilitation.
- Improved standardisation of assessment and documentation.
- Enhanced integration between inpatient and community pathways.
- Increased AHP confidence and cross-professional collaboration.
- Contribution to national learning and future BEPOP waves.

## Dissemination

Results and learning will be shared through:

- CPFT AHP Leadership and Quality Improvement forums.
- BEPOP national learning network.
- AGILE CSP Conference and BGS Age and Ageing meetings.
- Publication in *BMJ Open Quality* (post-implementation report) using the SQUIRE 2.0 guidance. (6)

A lay summary will be produced for CPFT Quality and Patient Experience reports.

## Funding

No external funding. This project will be conducted as part of routine quality improvement activity within CPFT.

## Conflict of Interest

None declared.

## Data Availability Statement

Data sharing not applicable as this work involves service-level quality improvement data collected within CPFT and reported in anonymised form through the national BEPOP framework.

## Author Contributions

I.S. conceived the project, drafted the protocol, and coordinated BEPOP integration within CPFT.

B.U. and A.M. provided methodological, clinical and operational oversight. All authors reviewed and approved the final manuscript.

## Acknowledgements

We acknowledge the support of the BEPOP Steering Group (NIHR Newcastle BRC, BGS & AGILE) and all the clinicians from CPFT contributing to this service improvement project.

## SQUIRE 2.0 Checklist

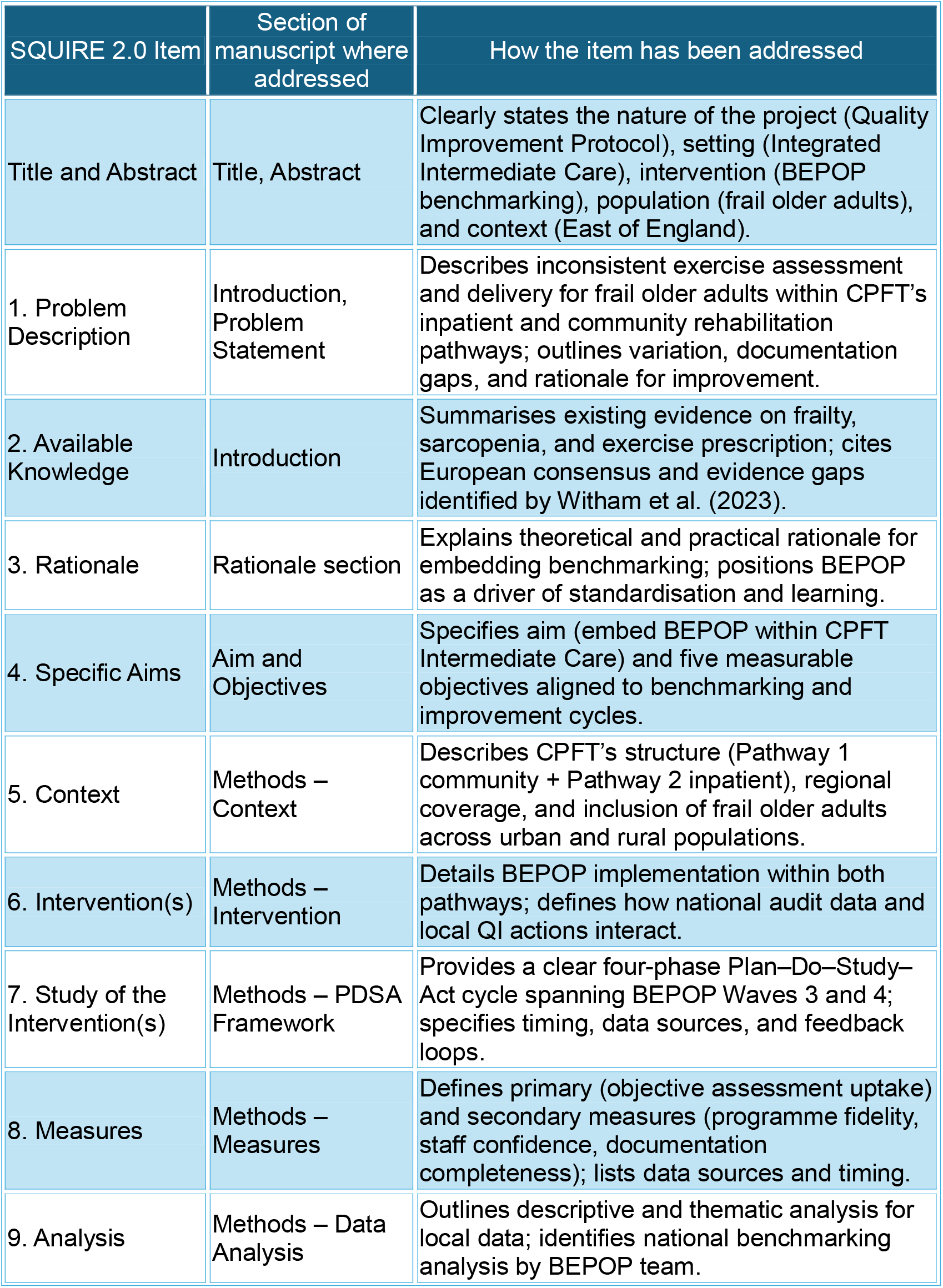
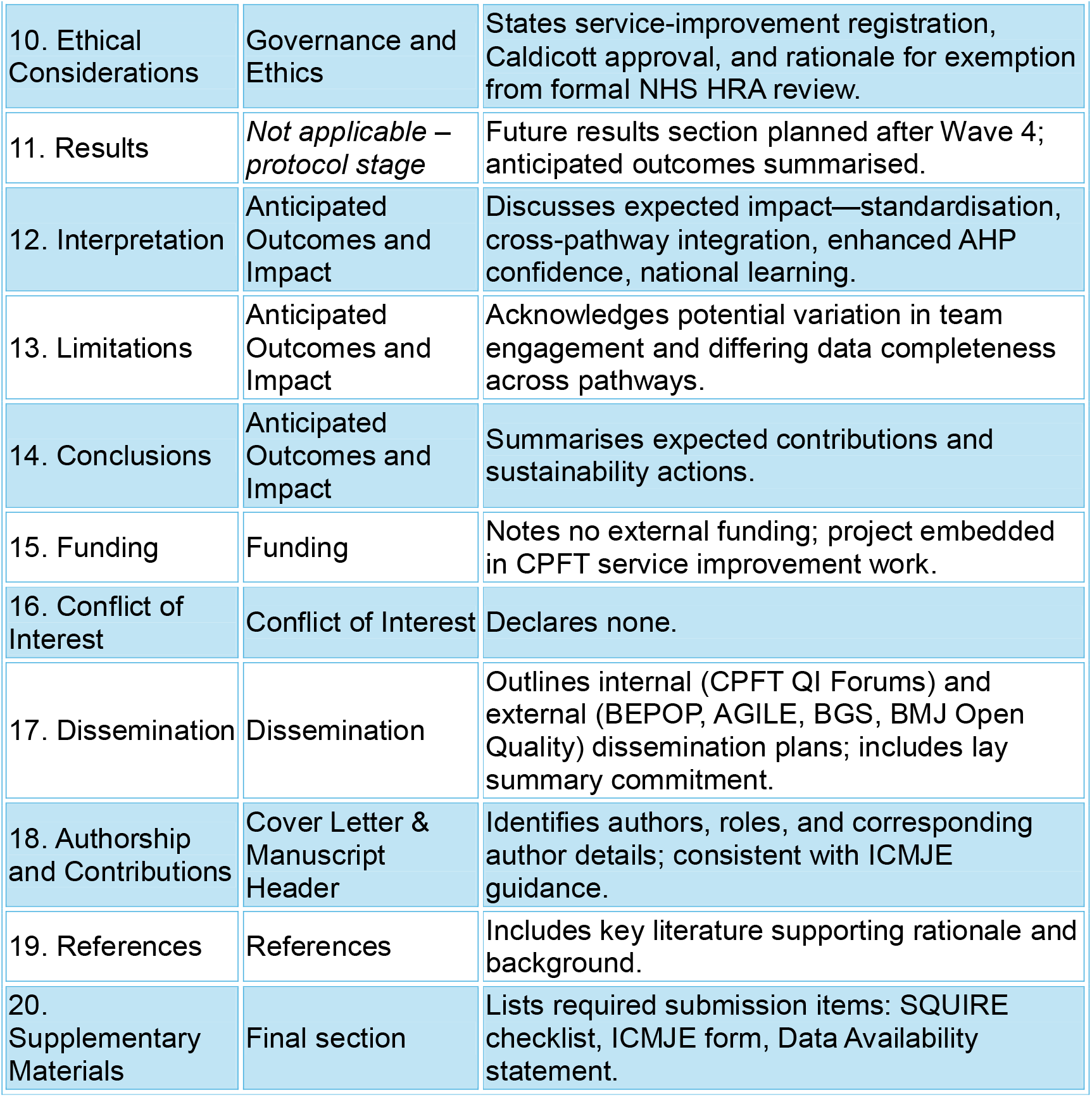

## Notes

### Competing Interest Statement

The authors have declared no competing interest.

### Funding Statement

No external funding received for this project. This project will be conducted as part of routine quality improvement activity.

### Author Declarations

Caldicott Guardian approval has been obtained for data sharing. As analysis involves anonymised routine care data, formal ethics review is not required for this service improvement project

